# Who cannot work from home? Characterizing occupations facing increased risk during the COVID-19 pandemic using 2018 BLS data

**DOI:** 10.1101/2020.03.21.20031336

**Authors:** Marissa G. Baker

## Abstract

**Objectives:** Not all workers are employed in occupations in which working from home is possible. These workers are at an increased risk for exposure to infectious disease during a pandemic event, and are more likely to experience events of job displacement and disruption during all types of public health emergencies. Here, I characterized which occupational sectors in the United States are most able to work from home during a public health emergency such as COVID-19.

**Methods:** 2018 national employment and wage data maintained by the U.S. Bureau of Labor Statistics (BLS) was merged with measures from the BLS O*NET survey data. The measures utilized rank the importance of using a computer at work, and the importance of working with or performing for the public, which relate to the ability to complete work at home.

**Results:** About 25% (35.6 M) of the U.S. workforce are employed in occupations which could be done from home, primarily in sectors such as technology, computer, management, administrative, financial, and engineering. The remaining 75% of U.S. workers (including healthcare, manufacturing, retail and food services, et al.) are employed in occupations where working from home would be difficult.

**Conclusions:** The majority of U.S. workers are employed in occupations that cannot be done at home, putting 108.4 M U.S. workers at increased risk for adverse health outcomes related to working during a public health emergency. These workers tend to be lower paid than workers who can work from home. During COVID-19, this could result in a large increase in the burden of mental health disorders in the U.S., in addition to increased cases of COVID-19 due to workplace transmission. Public health guidance to “work from home” is not applicable to the majority of the U.S. workforce, emphasizing the need for additional guidance for workers during public health emergencies.

## Introduction

Initial public health guidance for workers during the 2019-2020 COVID-19 pandemic was focused on ensuring workers stay home when sick, minimize non-essential travel, and practice good hygiene in order to slow the transmission of SARS-CoV-2 between workers and community members ^1^. As the number of cases grew, workers were urged or required to work from home, ^2,3^ schooling was moved online ^4,5^, retail establishments closed or severely reduced hours ^6,7^, and food establishments closed or moved to a model of takeout and delivery only ^8,9^. These measures, while necessary for halting the spread of a global pandemic, can have drastic effects on workers.

Exposure to infectious disease is often the primary consideration for worker health during a pandemic, particularly for front line workers such as those in healthcare. Previously, we calculated the number of workers in occupations where exposure to infection or disease occurs frequently using 2018 U.S. BLS occupational employment and O*NET data ^10^. We found that about 18% of the workforce is exposed to disease or infection at least once a month at work, putting these workers at an increased risk of not only contracting a disease due to work, but also transmitting an infectious disease into the community.

While exposure to infectious disease is an important occupational health concern during a pandemic, exposure to job insecurity (that is, concern about having a job in the future) is another important metric of worker health to consider. Several researchers have shown a relationship between exposure to acute and chronic job insecurity and measures of adverse physical and mental health outcomes including depression, stress, and physiologic markers such as increased blood pressure ^11–13^. Exposure to a job displacement event, due to voluntary or involuntary job loss stemming from a layoff, downsizing, or plant closure, also has been shown to be related to a variety of adverse mental health outcomes including depression, suicide, and stress ^14–16^, negative changes in diet ^17,18^, and physical health outcomes such as coronary heart disease and other physiologic markers of adverse health ^19,20^. After exposure to a job displacement event, workers may take jobs of lower quality, resulting in long-term economic and psychological effects for once-displaced workers ^21^. With many workers in the U.S. receiving healthcare and other benefits from their work arrangement, a layoff or reduction in hours can affect access to healthcare or long term stability for these workers ^22^.

Working from home can allow continued productivity when access to a workplace is restricted, such as during the COVID-19 pandemic. However, it is known that not all workers are able to work from home due to differences in job tasks. Jobs that lend themselves to being completed at home are jobs that require limited interaction with the public, so the work can be done without relying on others. Jobs that primarily use a computer to complete tasks also lend themselves to being done at home, given the portability of work on laptop computers.

When access to a workplace is restricted due to a public health emergency, the workers who cannot work from home are likely to experience job disruption, hours reduction, or voluntary or involuntary layoff. During COVID-19, this was exhibited fairly early, with joblessness claims in the U.S. hitting record highs, especially in occupations such as food service, retail, hospitality, and manufacturing ^23^. Further, workers who are essential personnel and continue going to workplaces (e.g. healthcare workers, grocery store workers, bus drivers) risk increased exposure to disease, and potential increases in job stress due to changes in job practices and duties to meet an increase in demand for services. The experiences of workers who cannot work from home will be different between occupations, informed by whether or not the work is essential, what workplace and regulatory protections exist for the occupation, the pay and benefits they receive, whether they have union protections, and how likely their industry is to return to normal operations after the pandemic event.

Here, I characterized which, and how many, United States workers perform job tasks that can be done at home, using metrics characterizing the importance of interacting with the public at work, and importance of computer use at work, and which groups of occupations are likely not able to work from home, putting them at risk for exposure to infectious disease at work, job displacement, disruption, or insecurity during this time. Additionally, I investigated how median annual wages differ between occupations that can and likely cannot work from home during a pandemic event, to better understand which workers may be most vulnerable to work disruptions during a pandemic event.

## Methods

This analysis utilized measures from two existing data sources, as previously detailed in Baker et al.^10^ and Doubleday et al.^24^. Briefly, U.S. employment and median annual wage by occupation, was downloaded from the U.S. BLS Occupational Employment Statistics database ^25^. These data were last updated in May 2018, and give a count of the number of U.S. workers employed in each 2010 Standard Occupational Classification code (2010 SOC) and the national median annual wage for each SOC. Guidance around SOC codes is detailed elsewhere ^26^ but briefly, SOC codes range from two digits (Major Group Code) to six digits (Detailed Occupation Code) and are hierarchical in nature. For example, SOC 35-0000 denotes “Food Preparation and Serving Related Occupations”, with SOC 35-9021 denoting the specific food preparation occupation of “Dishwasher.” For this analysis, six-digit occupation codes were utilized, and then aggregated over larger occupational groupings (i.e. two-digit codes).

To estimate the number of workers in occupations that could be done at home, the O*NET database was utilized. O*NET is a survey overseen by BLS that asks employees, employers, and job experts across six-digit SOC codes about exposures encountered at work, knowledge and skills utilized in the occupation, types of tasks performed, and workplace characteristics ^27^. O*NET does not collect data from military occupations; thus, SOC codes beginning with 55 “Military Specific Occupations” are not included in O*NET data. Similarly, employment numbers for “Military Specific Occupations” is not reported in the BLS Occupational Employment Statistics Database. All other SOC codes are included in the O*NET database, with updates made every year to ensure the database is completely refreshed every few years ^28^. Over a ten-year period (2001 to 2011) over 150,000 employees and job experts representing 125,000 workplaces had responded to the O*NET questionnaire, making it a robust source of occupational information ^29^.

Two O*NET measures were utilized in this analysis. The first characterized the importance of computer use at work via the question, “How important is working with computers to the performance of your current job?”. The second O*NET measure utilized was, “How important is performing for or working directly with the public to the performance of your current job?” For both questions, respondents could select from: Not Important, Somewhat Important, Important, Very Important, Extremely Important. Answers were converted to a 0-100 score, representing weighted-average score for each SOC code. A score of 50 is equivalent to a respondent answering “Important”.

Importance scores for both O*NET metrics were merged by six-digit SOC code with the national employment and annual median wage data. Annual median wage was used as opposed to annual mean wage in order to minimize effects from extreme values. Both O*NET measures were plotted against each other, with the resultant scatterplot divided into four quadrants. Each SOC on the scatterplot was weighted by annual median wage, to visualize differences in income between the four quadrants.

To further explore relationships in these data, the distribution of median annual wages was compared between quadrants using a Kruskall-Wallis test.

All data analysis was conducted using the statistical software package R version 3.6.3.

## Results

BLS reported a total of 144.7 million persons employed in the United States in May 2018; this does not include workers in military occupations. Figure 1 shows the relationship between “Importance of computer use at work” and “Importance of interaction with or performing for the public at work” for all 6-digit SOC codes. Each SOC plotted here is sized in proportion to the national median annual wage reported for that occupation by BLS, with larger points denoting a higher median annual wage. Each SOC on the plot is color-coded broadly by occupational sector.

**Figure 1:**
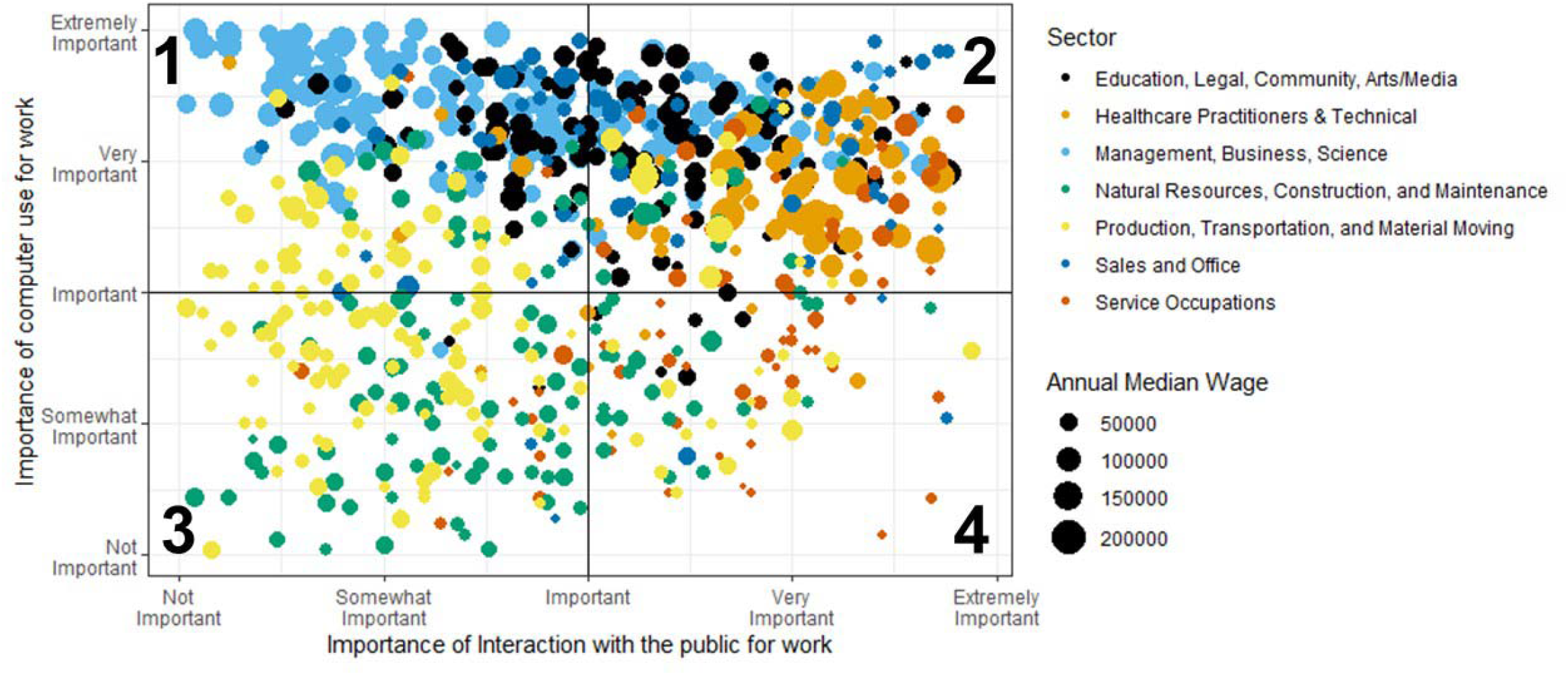
Who can work from home? Workers in quadrant 1 are workers that are likely able to work from home, whereas those in quadrants 2, 3, and 4 likely would not be able to work from home. Each point on the graph is weighted by the annual median wage for the occupation, and color-coded by broad occupational sector.

Figure 1 is divided into four numbered quadrants. SOCs in quadrant 1 represent those occupations that could likely be completed at home, that is, computer use is important to the work, but interaction with the public is not important. As detailed in Table 1, this quadrant represents 24.6% (35.6 M) of the BLS workforce and primarily includes occupational sectors such as business and finance, computer and mathematical, architecture and engineering, and the sciences, as shown in Table 2.

**Table 1:**
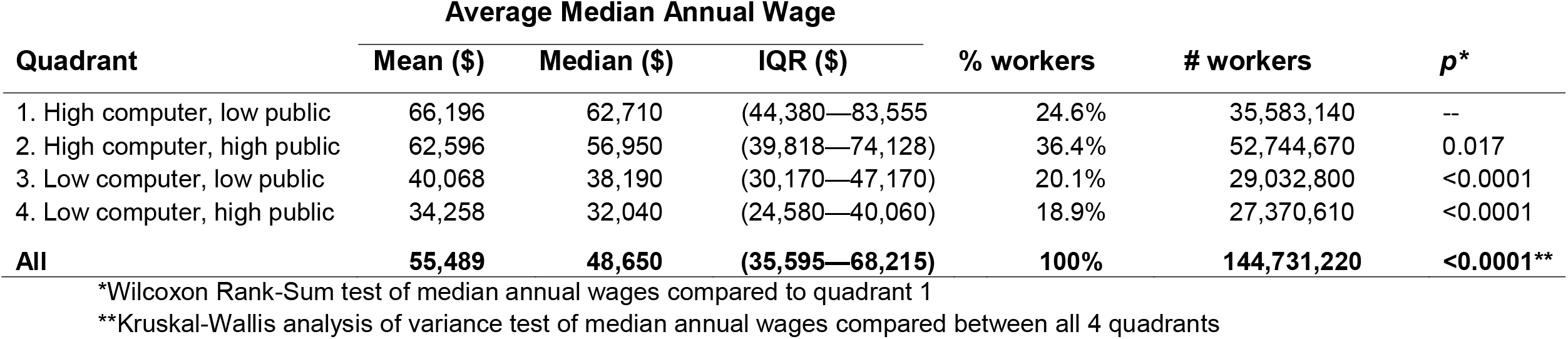
Distribution of median annual wages by quadrant, as shown in Figure 1

**Table 2:** Employment and annual average median wage by 2-digit SOC, and percentage of total SOC distribute across all four quadrants.

|  |  |  |  | % SOC distributed in each Quadrant |  |  |  |
| --- | --- | --- | --- | --- | --- | --- | --- |
| 2-digit SOC |  | Median Annual Wage | Total in SOC | 1 | 2 | 3 | 4 |
| 11 | Management | \$104,240 | 7,616,650 | 32.0% | 68.0% | -- | -- |
| 13 | Business & Financial Operations | \$68,350 | 7,721,300 | 72.7% | 27.3% | <0.1% | -- |
| 15 | Computer & Mathematical | \$86,340 | 4,384,300 | 100.0% | -- | -- | -- |
| 17 | Architecture & Engineering | \$80,170 | 2,556,220 | 90.8% | 9.2% | -- | -- |
| 19 | Life, Physical, & Social Science | \$66,070 | 1,171,910 | 63.6% | 36.3% | -- | -- |
| 21 | Community & Social Services | \$44,960 | 2,171,820 | -- | 97.8% | -- | 2.2% |
| 23 | Legal | \$80,810 | 1,127,900 | 9.9% | 90.1% | -- | -- |
| 25 | Education, Training, & Library | \$49,700 | 8,779,780 | 26.7% | 50.6% | 20.0% | 2.8% |
| 27 | Arts, Design, Entertainment, Sports, & Media | \$49,290 | 1,951,170 | 43.7% | 47.0% | -- | 9.3% |
| 29 | Healthcare Practitioners & Technical | \$66,440 | 8,646,730 | 7.7% | 90.5% | -- | 1.9% |
| 31 | Healthcare Support | \$29,740 | 4,117,450 | 1.3% | 38.7% | 36.6% | 23.3% |
| 33 | Protective Service | \$40,640 | 3,437,410 | 4.4% | 54.8% | -- | 40.8% |
| 35 | Food Preparation & Serving Related | \$23,070 | 13,374,620 | -- | 8.2% | 10.4% | 81.4% |
| 37 | Building & Grounds Cleaning & Maintenance | \$26,840 | 4,421,980 | -- | 2.3% | 70.4% | 27.3% |
| 39 | Personal Care & Service | \$24,420 | 5,451,330 | -- | 22.6% | 40.8% | 36.6% |
| 41 | Sales & Related | \$28,180 | 14,542,290 | 12.5% | 61.9% | 0.0% | 25.6% |
| 43 | Office & Administrative Support | \$35,760 | 21,828,990 | 45.6% | 52.0% | 0.8% | 1.6% |
| 45 | Farming, Fishing, & Forestry | \$25,380 | 480,130 | -- | 5.0% | 86.7% | 7.9% |
| 47 | Construction & Extraction | \$46,010 | 5,962,640 | -- | 2.2% | 76.0% | 21.9% |
| 49 | Installation, Maintenance, & Repair | \$45,540 | 5,628,880 | 30.7% | 23.3% | 38.9% | 7.0% |
| 51 | Production | \$35,070 | 9,115,530 | 21.1% | 1.7% | 74.2% | 3.0% |
| 53 | Transportation & Material Moving | \$32,730 | 10,244,260 | 4.8% | 5.9% | 48.4% | 41.1% |
| All SOCs | | \$38,640 | 144,733,290 | 24.6% | 36.4% | 20.1% | 18.9% |

**Bold** denotes the quadrant where the majority of workers in each 2-digit SOC fall.

The remaining three quadrants in Figure 1 represent occupations that likely cannot be done from home, making them susceptible to not only infectious disease exposure at work, but also to job disruption, job insecurity, and potential job displacement if their workplace closes. Quadrant 4 represents occupations where computer work is not important, and interaction with the public is very important. As detailed in Table 1, this quadrant represents 18.9% of the BLS workforce (27.4 M workers) and as shown on Table 2, consists of occupational sectors such as retail, food service, beauty services (e.g. barbers, hairdressers, manicurists), some protective services (e.g. security guards, TSA agents), and transportation operators such as bus drivers or subway operators.

Quadrant 2 represents occupations where both interaction with the public and computer use are important, and accounts for 36.4% (52.7 M) of the BLS workforce. These workers are in management, healthcare, legal, and education sectors. Quadrant 3 represents occupations where both interaction with the public and computer use are not important, accounting for 20.1% (29.0 M) of the BLS workforce. These are typically workers in construction, maintenance, production, and natural resources.

Table 1 summarizes the distribution of median annual wages across each quadrant. A Kruskall-Wallis one way analysis of variance test indicated that the median annual wage between these quadrants were significantly different. The quadrant with the highest annual median wage was quadrant 1, workers who could likely work from home, with a median annual wage of $62,710. The lowest annual median wage was in quadrant 4, which was $32,040. This over $30,000 difference is statistically significant when applying a Wilcoxon Rank-Sum test (p<0.0001).

Table 2 details total employment and median annual wage by 2-digit SOC, and the percentage of workers in each 2-digit SOC code that fall into each of the four quadrants.

## Discussion

During the COVID-19 pandemic, U.S. workers were urged or required to work from home help halt disease transmission. However, only about a quarter of U.S. workers are in occupations that can be done at home, with about 75% of the U.S. workforce (represented in Quadrants 2-4 on Figure 1) either remaining in the workplace and risking increased exposure to infectious disease, or experiencing job insecurity, disruption, and displacement due to workplace closure. The occupations that can be done at home have, on average, higher median wages than occupations that cannot be done at home, further increasing the vulnerabilities between these two groups.

Differences in exposure and experiences for these three quadrants that cannot work from home must be noted. Those workers in quadrant 2 are workers in jobs where interaction with both the public and computers are important. Many of these workers are in essential services, such as healthcare and education, making them less likely to be displaced from work, but still experience work disruptions due working different hours, performing different tasks, or working in a new modality. The workers that continue to go to work will also face increased exposure to disease. Like the workers in quadrant 1, these workers tend to have wages above the national median, and likely increased access to benefits and job protections through union and workplace protections.

Those in quadrant 3 are largely in construction, maintenance, natural resources, and manufacturing. Many of these workers are in jobs that may not be considered to be essential services ^30^ making them susceptible to job displacement or hours reductions if a shelter-in-place is ordered. If workplaces are open, workers may work in close proximity to other workers on jobsites, increasing risk of exposure to disease. Despite lower than median wages, many of these workers may have some protections from their union in addition to other regulatory protections, and increased certainty of a return to work when public health orders are lifted, given the vital nature of their work.

The workers in quadrant 4 are those workers for which using a computer is not important, but interacting with the public is. These workers, largely in food services, some protective services, personal care, and transportation could face job displacement and job insecurity as nonessential business are asked to close for public health reasons, and the public avoids nonessential activities. Those working in grocery stores and other essential retail be less likely to face job displacement during a public health emergency as their workplaces will remain open. However, if schools close, essential workers that cannot work from home may have to choose between quitting their job or reducing their hours in order to stay home with children, or going into work without adequate back-up care for their children, further contributing to a feeling of insecure employment and stress. Importantly, these workers are also at increased risk of exposure to SARS-CoV-2, and may have to choose between continuing to work and risking exposure, or quitting with no safety net, which could be a particularly challenging decision for a worker in a high-risk group (e.g. older, pregnant, immunocompromised).

Other data sources have quantified the number of U.S. workers that work from home, including the American Time Use Survey, U.S. Census American Community Study, and the National Compensation Survey. However, these data sources do not quantify how many and which types of workers have work that feasibly can be done at home when workers are ordered to do so. The National Compensation Survey characterizes how many workers have access to a remote working benefit as part of a compensation package, regardless of whether a respondent took advantage of it ^31^, and The American Time Use Survey and the American Community Study characterize whether a worker worked from home on the day the survey was administered, regardless of whether it was paid work or not ^32,33^. The work presented here quantified the number and types of workers who could work from if it was ordered in an emergency, which is an important distinction from the above-mentioned data sources.

The COVID-19 pandemic, and other public health emergencies and disasters, tend to exacerbate existing disparities in society, which was also shown in this analysis. Here, I showed that the distribution of median annual wages differed between those workers that would likely be able to work from home, and those workers that would likely not be able to work from home, further adding to the vulnerability of lower-income workers during the COVID-19 pandemic. Workers who are able to work from home will have some continuity in pay, increased ability to care for a child out of school, decreased risk of being laid off or having hours substantially cut, and decreased potential exposure to disease or infection via other workers or community members. This further exemplifies the importance of work as a social determinant of health, and highlights the importance of understanding which workers are in more vulnerable jobs during an emergency or disaster, and the risks and challenges these groups face ^34^.

Limitations related to the data used here must be acknowledged. BLS data does not count self-employed (which includes a variety of workers ranging from gig economy workers to highly trained independent consultants, for example), undocumented, contingent, military, and domestic workers. This undercoverage of the working population in the BLS survey could affect conclusions presented here. O*NET relies on employee and employer self-report, so is subject to inherent bias and misclassification during collection. Further, data collected by O*NET is aggregated on the occupational level, and I further aggregated data into quadrants, meaning that within-occupation and within-quadrant variation isn’t accounted for in this analysis.^35^ This will lead to misclassification both within the occupations, and within each quadrant. The O*NET metrics used in this analysis were measures of the importance of using a computer for work and importance of interacting with the public, which differs from the frequency of using a computer or frequency of interacting with the public. Therefore, some jobs for which computer use is rated as very important, may not actually require use of a computer very frequently, and jobs where interaction with the public is rated as important may not actually interact with the public frequently. This further lead to misclassification in the analysis for who could work from home most easily.

## Public Health Implications

Understanding the unique challenges that workers who cannot work from home could face during a pandemic or other public health emergency can help to inform appropriate risk management and policy-based strategies for these workers, to ensure that their livelihood can continue. This work shows that only about 25% of the United States workforce are in jobs that could continue to be done at home during a pandemic event. These workers would be protected from disease exposure due to working from home, and are typically higher paid jobs with more workplace protections, further protecting these workers from adverse health effects related to job insecurity, job stress, and job displacement.

The rest of the workforce (about 75% across quadrants 2-4 in Figure 1) are in occupations that would face increased exposure to disease and infection if they are still working out of their workplaces, and increased exposure to psychosocial factors such as job displacement, disruption, and insecurity if they are not able to work out of their workplaces. These workers could also face stress and job insecurity as they may have to choose between going to work and being exposed, and staying home to protect themselves or care for a family member.

Experiences and outcomes for these workers during a pandemic event would likely be modified by workplace characteristics, such as available workplace controls, workplace policies and benefits, whether or not workers are unionized, whether or not workers qualify for state or federal unemployment protections, worker pay, and the probability of returning to work once normal operations resume.

In this analysis, I found that the workers with the lowest average median wage are workers that are not able to work from home, and include occupational groups such as food services, retail, personal care, and some transportation workers. Workers in this quadrant have an average annual median wage about $30,000 less than the workers who can work from home, and these workers often lack protections such as employer-provided healthcare, appropriate sick leave, or paid time off, further increasing their vulnerability during a public health emergency, and enforcing the role of work as a social determinant of health.

While all workers will be disrupted during a pandemic event such as COVID-19, increased public health focus should be on those that are the most vulnerable, including ensuring these workers are adequately protected at work, and have social protections in the event they no longer are able to work. This will ensure these workers do not bear an undue health burden during a public health emergency, and also help to reduce the burden of adverse health outcomes that could emerge in these workers who cannot work from home during a pandemic event.

## Data Availability

All data used in this analysis is publicly available through the US Bureau of Labor Statistics, with links referenced in the manuscript.

https://www.bls.gov/oes/home.htm

https://www.onetonline.org/

## Notes

### Competing Interest Statement

The authors have declared no competing interest.

### Funding Statement

Research reported here was supported by the National Institute for Occupational Safety and Health under Federal Training Grant T42OH008433. The content is solely the responsibility of the authors and does not necessarily represent the official views of NIOSH.

